# Myocardial Native T1 Mapping in the German National Cohort (NAKO): Associations with Age, Sex, and Cardiometabolic Risk Factors

**DOI:** 10.1101/2025.07.16.25331651

**Authors:** Clemens Ammann, Jan Gröschel, Hadil Saad, Susanne Rospleszcz, Christopher Schuppert, Thomas Hadler, Richard Hickstein, Thoralf Niendorf, Janis M. Nolde, Matthias B. Schulze, Karin H. Greiser, Hermann Brenner, Ute Mons, Josua A. Decker, Thomas Kröncke, Thomas Küstner, Konstantin Nikolaou, Stefan N. Willich, Thomas Keil, Marcus Dörr, Robin Bülow, Fabian Bamberg, Tobias Pischon, Christopher L. Schlett, Jeanette Schulz-Menger

## Abstract

**Background:** In cardiovascular magnetic resonance (CMR), myocardial native T1 mapping enables quantitative, non-invasive tissue characterization and is sensitive to subclinical changes in myocardial structure and composition. However, data on the association between cardiometabolic risk and myocardial alterations are still limited. We therefore investigated how age, sex, and cardiometabolic risk factors are associated with myocardial T1 as a potential imaging marker of myocardial target-organ involvement in a population-based analysis within the German National Cohort (NAKO).

**Methods:** This cross-sectional study included 29,573 prospectively enrolled participants who underwent midventricular T1 mapping using 3.0 T CMR along with deep clinical phenotyping. After artificial intelligence-assisted myocardial segmentation, a subset of 9,162 outlier cases was subjected to manual quality control according to clinical evaluation standards. Cardiometabolic risk factors were identified through self-reported, physician-diagnosed medical history, clinical chemistry, and blood pressure measurements. Associations with myocardial T1 were evaluated using multiple linear regression models adjusted for age, sex, heart rate, imaging site, and comorbidities.

**Results:** After quality control, 27,794 participants (44.6% women; 20–75 years) were included. Mean T1 was higher in women (1,231 ± 33 ms) than in men (1,209 ± 34 ms), with sex differences progressively declining with age. In adjusted analyses, T1 was significantly higher in individuals with diabetes, kidney disease, and current smoking. Conversely, hyperlipidaemia was significantly associated with lower T1. Associations with hypertension showed strongly sex-specific patterns: women had lower T1 values, while T1 increased with hypertension severity in men.

**Conclusions:** Myocardial native T1 varies by sex and age and shows distinct sex-specific associations with major cardiometabolic risk factors, supporting its role as a sensitive imaging marker of subtle myocardial alterations. Unexpectedly lower T1 times in participants with hyperlipidaemia suggest a potential direct effect of blood lipids on the heart, which warrants further investigation.

Graphical Abstract
Associations of Age, Sex, and Cardiometabolic Risk Factors with Myocardial Native T1 in the NAKO Study
Bottom left panel: unstandardized β coefficients and 95% confidence intervals from multiple linear regressions, stratified by sex. Bottom right panel: linear regression (solid line), median (dotted line), and interquartile range (shaded area) in 2-year age bins, stratified by sex. The last bin is defined as ≥ 70 years. MOLLI: modified look-locker inversion recovery; LDL-c: LDL-cholesterol, AI: artificial intelligence; SD: standard deviation; HR: heart rate; CVD: cardiovascular disease; TD: thyroid disease; IQR: interquartile range

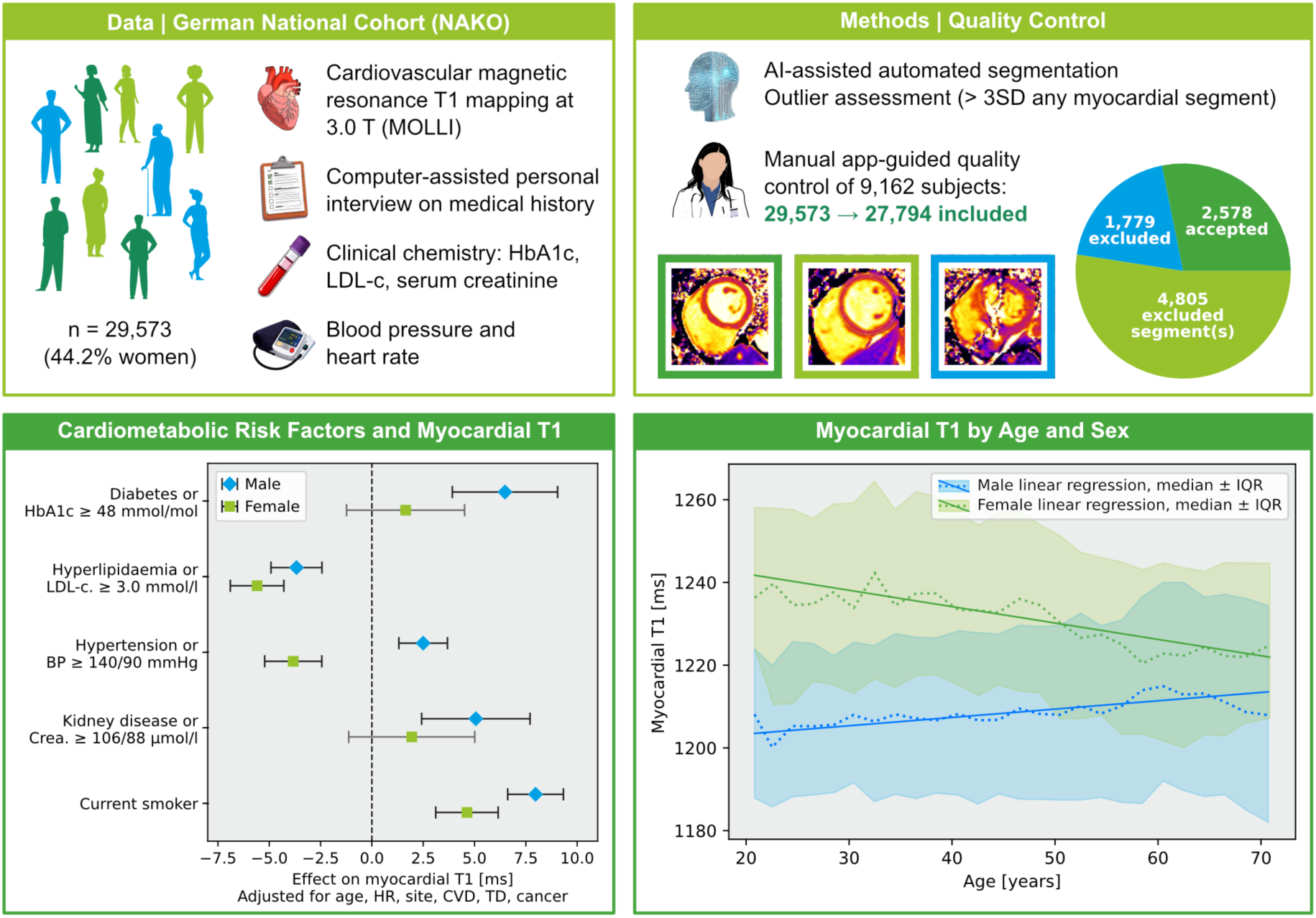

## Introduction

Myocardial native T1 mapping by cardiovascular magnetic resonance (CMR) is a quantitative, non-invasive imaging marker of myocardial tissue composition [1,2]. The technique is sensitive to diffuse processes including inflammation, oedema, interstitial fibrosis or myocardial infiltration with iron or lipid deposits and has proven helpful in diagnosing and monitoring cardiovascular disease [3]. Importantly, T1 mapping enables the detection of subclinical changes in myocardial tissue composition, potentially capturing early stages of cardiac pathology [4].

The German National Cohort (NAKO) is a population-based prospective study comprising over 100,000 women and 100,000 men aged 19 to 75 years with extensive phenotyping, aimed at investigating the causes of chronic diseases [5]. Its whole-body magnetic resonance (MR) imaging subcohort comprises 30,868 individuals who underwent standardized 3.0 T MR imaging at five centres across Germany with near-simultaneous clinical and laboratory assessments [6]. Given the high prevalence of cardiometabolic comorbidities and metabolic syndrome in the general population and the NAKO study [7], non-contrast CMR offers a promising tool for detecting myocardial alterations in these individuals.

Major cardiometabolic risk factors such as diabetes, hypertension, hyperlipidaemia, chronic kidney disease, and smoking are believed to promote adverse structural myocardial remodelling through mechanisms including low-grade inflammation and fibrosis [8–12]. Native T1 mapping may therefore serve as a sensitive, albeit non-specific, imaging marker of myocardial target-organ involvement. In the UK Biobank, T1 mapping at 1.5 T was found to be associated with prevalent cardiovascular disease and incident events in participants aged 44 to 84 based on hospital registry data [13]. However, large-scale prospective studies examining sex-stratified effects of cardiometabolic risk factors on myocardial tissue composition are still lacking, and data from younger individuals and from 3.0 T imaging remain limited. Moreover, while previous studies have reported sex differences of myocardial native T1, the role of age remains inconsistent and insufficiently understood [13–15].

In this cross-sectional analysis of the NAKO MR cohort, we investigated how cardiometabolic risk factors, age, and sex are associated with native T1 as a marker of myocardial tissue composition.

## Methods

### Study Population

This study leverages data from the NAKO at baseline [6] and included 29,573 participants (44.2% women) with a midventricular T1 map obtained by CMR and corresponding data from self-questionnaires and clinical chemistry. Participants in the NAKO MR cohort were randomly selected from resident registers and scanned between 2014 and 2019 across five German imaging centres, ensuring uniform protocols and equipment (3.0 T MAGNETOM Skyra, Siemens Healthineers, Forchheim, Germany). A modified look-locker inversion recovery (MOLLI) 5(3)3 T1 mapping technique was used to acquire a midventricular short-axis T1 pixel map. Imaging parameters are published by Bamberg et al. [16] and include a voxel size of 1.4×1.4×8.0 mm and a flip angle of 35°. The average interval between clinical examination and MR imaging was 28 to 42 days.

### Analysis and Quality Control

Automatic segmentation of the myocardium and identification of the right ventricular insertion point were performed using a deep learning-based research algorithm (Siemens Healthineers, Forchheim, Germany) [17,18]. Mean T1 values were calculated globally and for the six midventricular segments defined by the American Heart Association (AHA) [19] using dedicated in-house research software [20].

After exclusion of repeated or failed T1 mapping acquisitions, image quality and artificial intelligence (AI)-generated segmentations were reviewed manually by an experienced physician if the mean T1 time of at least one myocardial segment differed by more than three standard deviations from age- (10-year groups) and sex-specific means. A custom Python tool enabled visual inspection of T1 maps with overlayed contours alongside corresponding source images and metadata. In case of insufficient image quality due to inadequate breathing motion correction or co-registration of individual images, electrocardiographic trigger errors, incorrect orientation or other image artifacts not limited to individual myocardial segments, the entire examination was excluded from analysis. If at most three segments had to be excluded, the mean myocardial T1 time was calculated over the remaining segments weighted by their area. Manual quality control followed recommendations for clinical evaluation [1].

To validate the analysis pipeline, 100 representative cases (matched for age, sex, height, weight, blood pressure as well as cardiovascular and metabolic condition prevalence) were reanalysed manually in CE-certified medical software and compared with results from automated segmentation followed by quality control.

### Statistical Analysis

Statistical analyses were carried out in Python 3.12 using statsmodels 0.14.4 and following recommendations for analysing NAKO data [21]. Variables of interest included: age, sex, body mass index (BMI), blood pressure, heart rate, self-reported cardiovascular or metabolic diseases, current smoking, and laboratory parameters (HbA1c, LDL-cholesterol, serum creatinine). Medical history was collected via standardized computer-assisted personal interview limited to physician-diagnosed conditions. Variables were primarily sourced from the clinical examination, with age extracted from DICOM metadata.

Mean T1 relaxation time was analysed by sex and myocardial segment. Since lateral segments generally exhibit more artifacts and lower T1 values, all statistical analyses were additionally performed for the two septal segments only and are reported in the Supplementary Material. A healthy subcohort was defined as follows: none of the diseases listed in Table 1, no current smoking, BMI between 18.5 and 30.0 kg/m^2^, blood pressure < 140/90 mmHg, LDL-cholesterol < 3.0 mmol/l (116 mg/dl), HbA1c < 48 mmol/mol (6.5%) and serum creatinine < 106 µmol/l (1.2 mg/dl) for men or < 88 µmol/l (1.0 mg/dl) for women. By combining reported conditions and clinical chemistry, we aimed to exclude both treated patients with controlled disease and undiagnosed diseased participants.

**Table 1:**
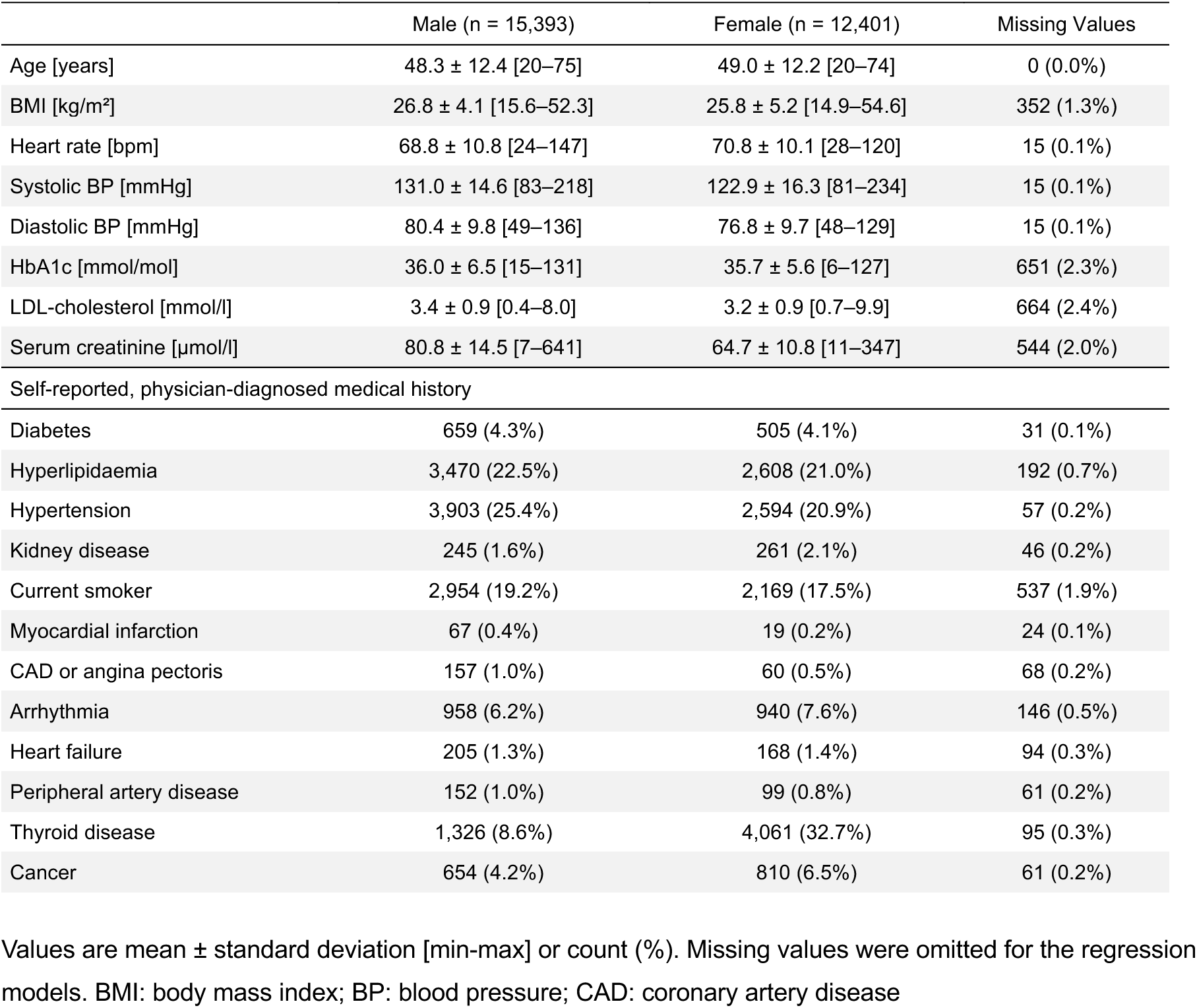
Basic Characteristics of Participants and Prevalence of Self-Reported Conditions.

Ordinary least squares linear regression was used to examine the associations of age, sex, and their interaction (age×sex) with myocardial T1 in the full cohort. Heart rate and imaging site were included as covariates due to the potential sensitivity of the MOLLI sequence to heart rate [1,22] and minor variations between sites (Supplementary Figure 1). Age was mean-centred to aid interpretation and reduce collinearity. Unstandardized regression coefficients β with 95% confidence intervals are reported to facilitate direct interpretation in the unit of T1 (ms). We observed a strong sex dependency that varied by age. This relationship was visualized using linear regression, median and interquartile range in 2-year bins. Heart rate was linearly associated with T1 (β = 0.35 [0.31–0.38] ms/bpm; *p* < 0.001). The five imaging sites revealed small but statistically significant differences (all *p* ≤ 0.008). Accordingly, all subsequent analyses were adjusted for heart rate, imaging site, age, sex, and their interaction; where appropriate, analyses were also stratified by sex.

To evaluate the association between cardiometabolic risk factors and myocardial T1 relaxation times, we used multiple linear regression models with T1 as the dependent variable. Independent variables of interest comprised the presence of self-reported diabetes or elevated HbA1c, reported hyperlipidaemia or elevated LDL-cholesterol, reported hypertension or elevated measured blood pressure, reported kidney disease or elevated creatinine levels, and current smoking. The limits for blood parameters corresponded to those defined for the healthy subcohort. For LDL-cholesterol, the target value for low-risk populations (< 3.0 mmol/l) specified by the European Society of Cardiology (ESC) and European Atherosclerosis Society was used [23]. Other cardiovascular disease (myocardial infarction, coronary artery disease, heart failure, arrythmia, peripheral artery disease) as well as thyroid disease and cancer were included as covariates in the regression model. Detailed medication data were not available in the full cohort and could therefore not be included as covariates; the reported associations thus reflect cardiometabolic risk independent of specific treatment effects.

In a sub-analysis, blood pressure was categorized according to the ESC hypertension classification [24] into grade I (≥ 140/90 mmHg), grade II (≥ 160/110 mmHg), grade III (≥ 180/110 mmHg), and isolated systolic or diastolic hypertension. We calculated sex-stratified linear regression models with myocardial T1 as the dependent variable and hypertension category as the main independent variable, adjusting for all other variables from the primary model. In a further linear regression analysis restricted to participants with self-reported hyperlipidaemia, the influence of lipid-lowering therapy was examined, adjusting for all covariates from the previous models.

Variance inflation factors remained below 2.0, indicating no relevant multicollinearity. Subjects with missing values were omitted from the respective analyses. BMI was excluded from the regression models as it may act as a mediator in the causal pathway from lifestyle or metabolic factors to cardiometabolic conditions such as hypertension and diabetes. Adjusting for BMI in this context could attenuate the observed effects of these conditions on myocardial T1.

Nonlinear associations between BMI, mean arterial pressure (MAP) and heart rate with myocardial T1 were explored separately using locally estimated scatterplot smoothing stratified by sex. Medians and 95% confidence intervals were bootstrapped for equal-width bins, and sex-stratified density plots were displayed along the same horizontal axis for each variable.

### Ethics

This study was approved by the NAKO Use and Access Committee, based on the participants’ informed consent and consistency with the objectives of the NAKO. Ethical approval for the NAKO study was obtained from the responsible local ethics committees of all study centres. The study was conducted in accordance with the Declaration of Helsinki and subsequent amendments.

## Results

### Quality Control

Of the 29,573 available T1 maps, 9,162 images with corresponding myocardial segmentation were selected for manual quality control, following the criteria outlined above. Among these, 1,779 subjects were excluded entirely from the analysis, while 4,805 cases involved the exclusion of one to three individual myocardial segments. The quality control procedure and results are illustrated in Figure 1. Artifacts were most frequently found in the inferolateral (n = 3,634) and inferior (n = 2,761) myocardial segments. The exclusions reduced the standard deviation of the overall mean T1 in the final dataset from 48 ms to 35 ms, while the mean itself remained unchanged (difference < 0.1 ms). Following quality control, a total of 27,794 (44.6% female) participants were included in the final analysis. Manual validation showed excellent agreement with automated results (intraclass correlation ICC(3,1) = 0.96 [0.94–0.97]; *p* < 0.001; Supplementary Figure 2).

**Figure 1:**
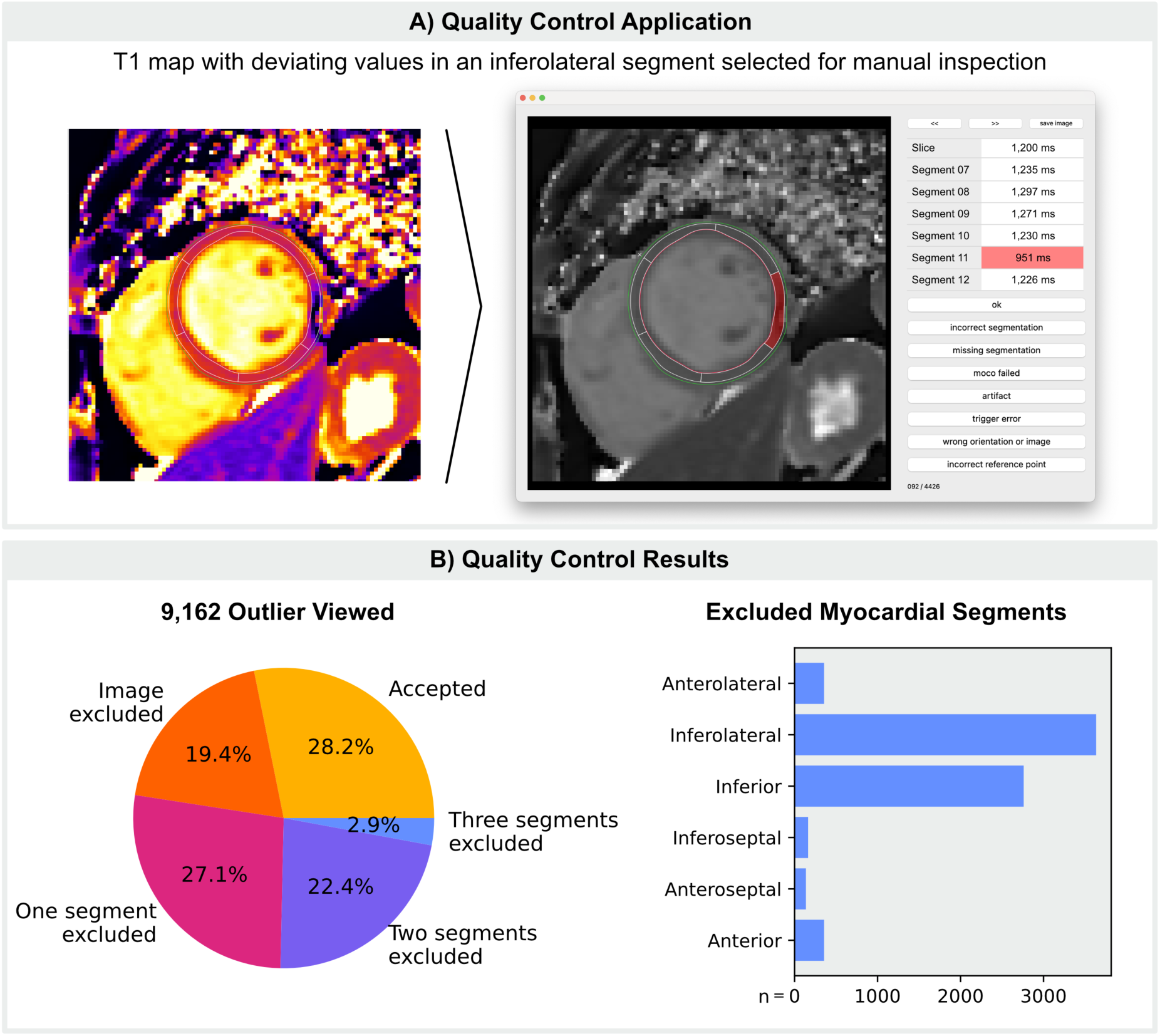
Manual Quality Control. A) T1 maps with abnormal values that deviate by more than three standard deviations from age- and sex-specific means in at least one myocardial segment are displayed together with the artificial intelligence-generated segmentation in a Python-based interface. Individual segments with artifacts can be excluded from the analysis by clicking on the respective area. B) Results from manual quality control.

### Participant Characteristics

Participant characteristics and the prevalence of self-reported conditions are summarized in Table 1. Following the definition outlined above, 3,910 (14.1%) participants were assigned to the healthy subcohort. For the combination of self-reported medical history and elevated blood parameters, 1,341 (4.8%) participants reported diabetes or had elevated HbA1c, 18,263 (65.7%) reported hyperlipidaemia or had elevated LDL-cholesterol, 10,363 (37.3%) reported hypertension or had elevated blood pressure, and 1,147 (4.1%) reported kidney disease or had elevated serum creatinine levels.

### Age and Sex Dependence of Myocardial T1

The mean myocardial T1 relaxation time for 27,794 participants was 1,219 ± 35 ms at 3.0 T. Septal T1 (1,237 ± 40 ms) was significantly (*p* < 0.001 by two-sided paired t-test) higher than in lateral segments (1,200 ± 46 ms). Inferolateral and inferior segments had the highest variance in T1 values (Figure 2).

**Figure 2:**
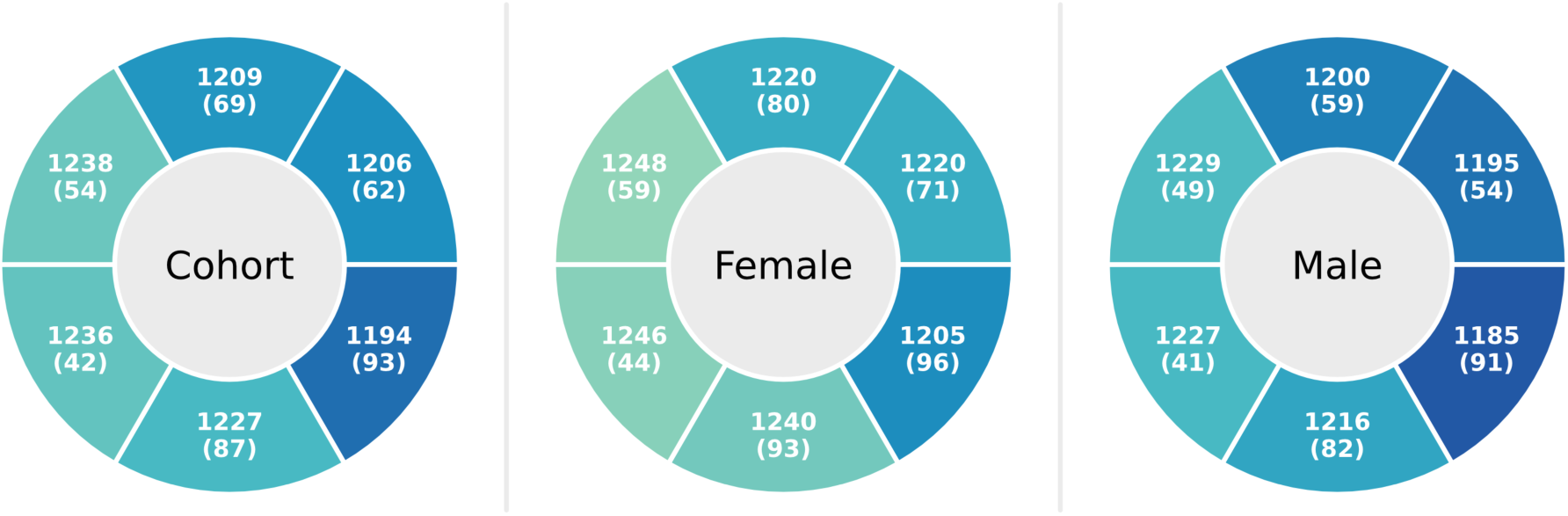
Polar Plots for Mean Myocardial T1 per AHA Segment. Values are given as the mean and average standard deviation of T1 within each segment (i.e., the mean of the standard deviations of individual pixel values). AHA: American Heart Association

In adjusted linear regression models, sex was significantly associated with myocardial T1 relaxation time (β = 20.96 [20.18–21.74] ms at the mean age of 48.6 years; *p* < 0.001). However, the effect of sex varied with age and showed a significant interaction (*p* < 0.001). While T1 increased with age in male participants (β = 0.20 [0.16–0.24] ms/year; *p* < 0.001), the age dependence was even stronger but inverse for women (β = −0.37 [−0.42 to −0.33] ms/year; *p* < 0.001) when stratified by sex. Visually, the age dependency appeared less pronounced in the healthy subcohort (Figure 3), which was statistically confirmed by a significant three-way interaction term (*p* = 0.009). Mean values are shown in Table 2 for 10-year age groups (with the last bin covering ages 60 to 75). Corresponding values for septal segments only are provided in Supplementary Table 1.

**Figure 3:**
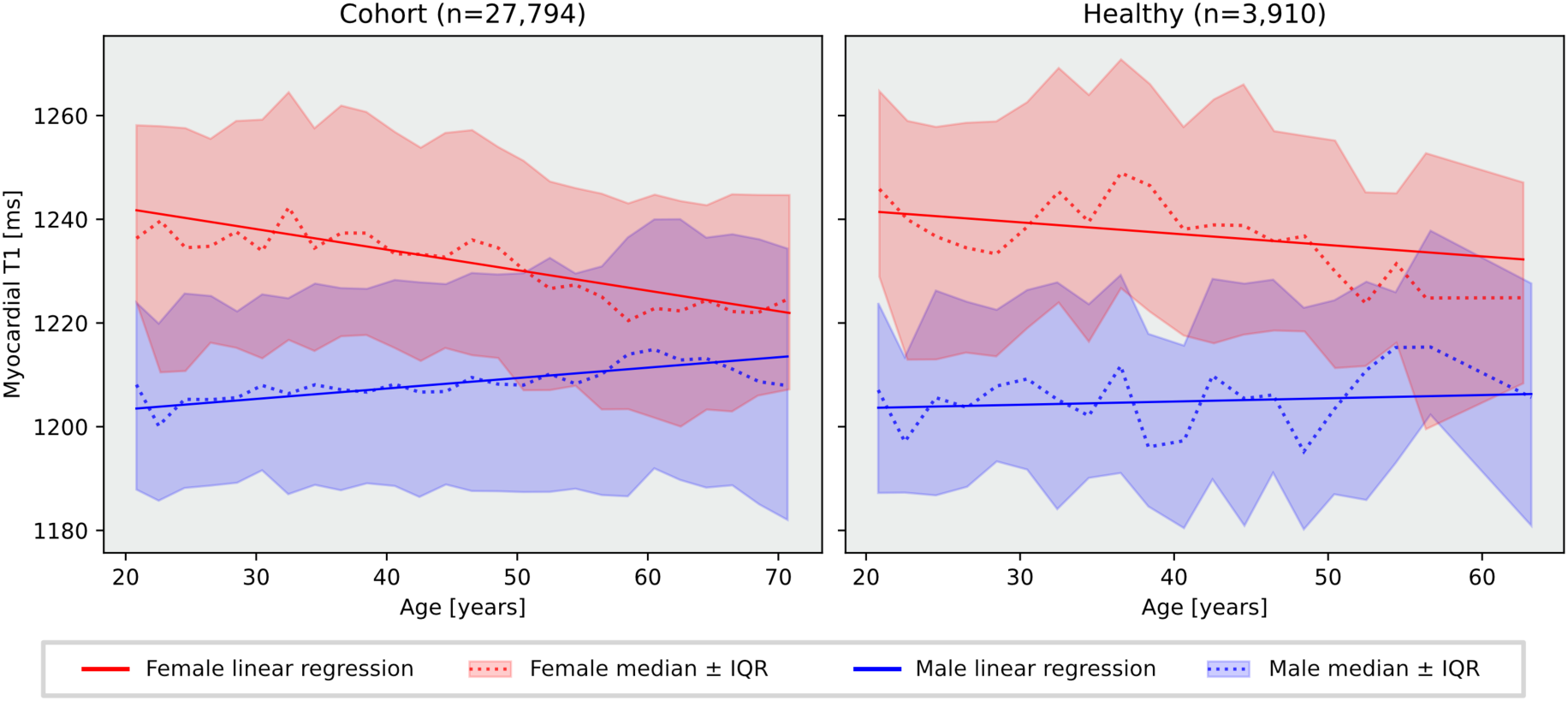
Age and Sex Dependence of Myocardial T1. Linear regression (solid line), median (dotted line), and interquartile range (shaded area) shown in 2-year age bins, stratified by sex. The last bin is defined as ≥ 70 years for the whole cohort and ≥ 58 years for the healthy subcohort.

**Table 2:** Mean Myocardial T1 for Age Groups by Sex.

| Age Group | Male |  | Female |  |
| --- | --- | --- | --- | --- |
|  | mean ± SD | n | mean ± SD | n |
| 20-29 | 1,205 ± 28 | 1,532 | 1,235 ± 33 | 1,167 |
| 30-39 | 1,207 ± 30 | 2,082 | 1,238 ± 34 | 1,427 |
| 40-49 | 1,208 ± 31 | 4,246 | 1,235 ± 32 | 3,445 |
| 50-59 | 1,210 ± 35 | 4,348 | 1,227 ± 31 | 3,643 |
| 60-75 | 1,213 ± 38 | 3,185 | 1,224 ± 32 | 2,719 |
| Total | 1,209 ± 34 | 15,393 | 1,231 ± 33 | 12,401 |

Two-way analysis of variance: age group $p < 0.001$ ; sex $p < 0.001$ ; age group-sex interaction $p < 0.001$ . SD: standard deviation; n: number of subjects

### Associations of Cardiometabolic Risk Factors with Myocardial T1

Multiple linear regression analyses adjusted for age, sex, age×sex, heart rate, imaging site, and relevant comorbidities yielded the following results: Self-reported diabetes or elevated HbA1c (β = 4.14 [2.23–6.06] ms; *p* < 0.001) and kidney disease or elevated serum creatinine levels (β = 3.64 [1.63–5.64] ms; *p* < 0.001) were both associated with significantly higher T1 times in the overall cohort. When stratified by sex, both associations remained significant in men (*p* < 0.001) but not in women (*p* = 0.263 and *p* = 0.215, respectively). Current smoking was associated with elevated myocardial T1 (β = 6.60 [5.59–7.62] ms), while hyperlipidaemia or elevated LDL-cholesterol was significantly associated with lower myocardial T1 values in the overall cohort (β = −4.69 [−5.59 to −3.79] ms) and in both sexes (all *p* < 0.001). Sex-specific effects were observed for reported hypertension or elevated blood pressure, with a significant positive β in men (β = 2.50 [1.31–3.68] ms; *p* < 0.001) and a significant negative β in women (β = −3.83 [−5.22 to −2.44] ms; *p* < 0.001). In the entire cohort, the association was not significant (*p* = 0.931). Corresponding regression coefficients and confidence intervals are illustrated in Figure 4 and provided in Supplementary Table 2. A confirmatory analysis restricted to septal segments rendered comparable results (Supplementary Figure 3). Among participants with hyperlipidaemia, lipid-lowering therapy was associated with a nonsignificant, small decrease in myocardial T1 (β = −0.92 [−2.99 to 1.14] ms; *p* = 0.381).

**Figure 4:**
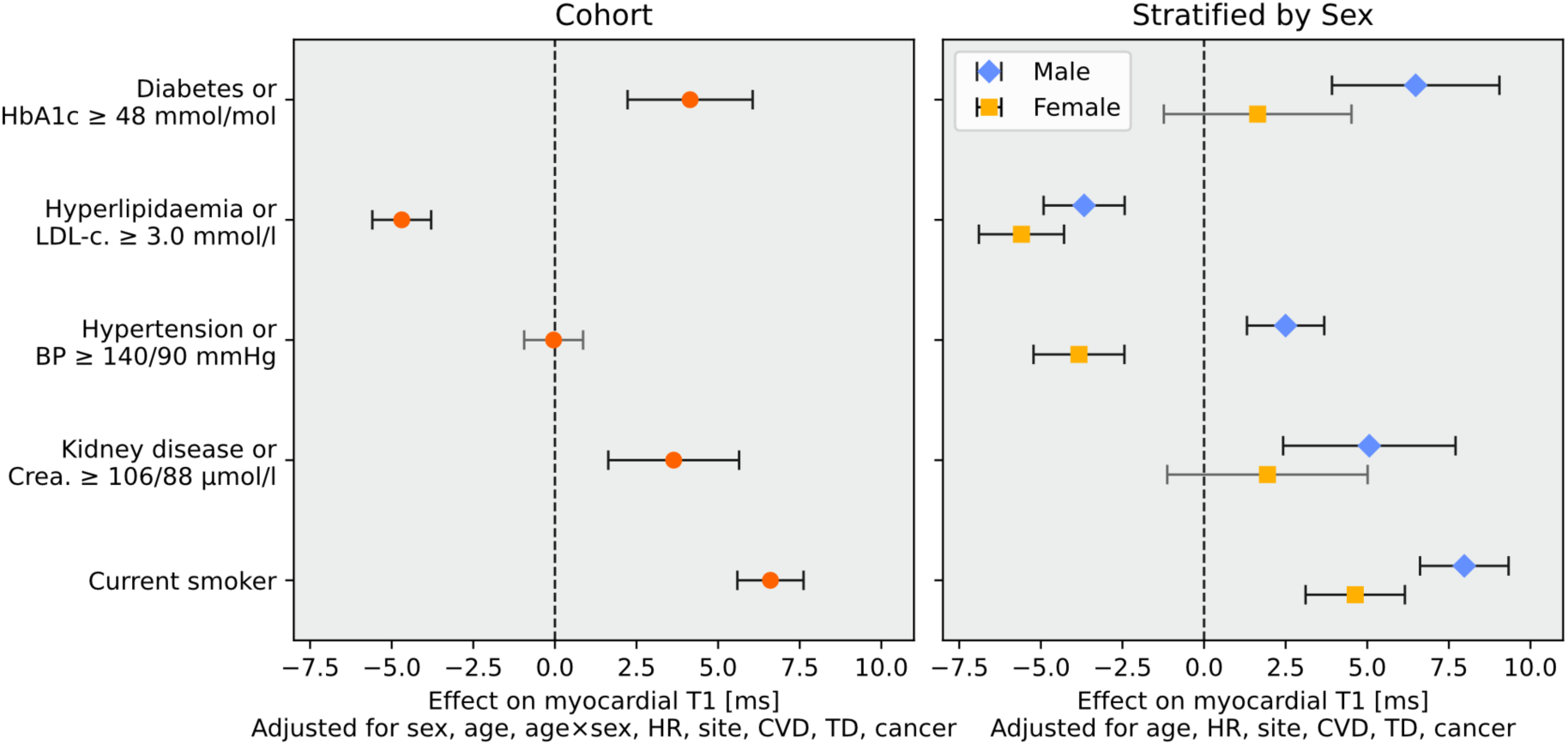
Associations of Cardiometabolic Risk Factors with Myocardial T1. Unstandardized β coefficients and 95% confidence intervals from adjusted multiple linear regressions for the whole sample (left panel) and stratified by sex (right panel). Covariates included sex, age, age×sex, heart rate (HR), imaging site, cardiovascular disease (CVD), thyroid disease (TD), and cancer. LDL-c.: LDL-cholesterol; BP: blood pressure; Crea.: serum creatinine

Body mass index exhibited a nonlinear, U-shaped relationship with myocardial T1, with higher T1 values observed in both underweight and overweight individuals (Figure 5). Consistent with the findings from the regression model, there was a pronounced and unexpected negative correlation at low blood pressure values (MAP < 90 mmHg) in female participants, while blood pressure was almost linearly positively correlated in male participants. The heart rate showed an approximately linear positive relationship to T1 for both sexes.

**Figure 5:**
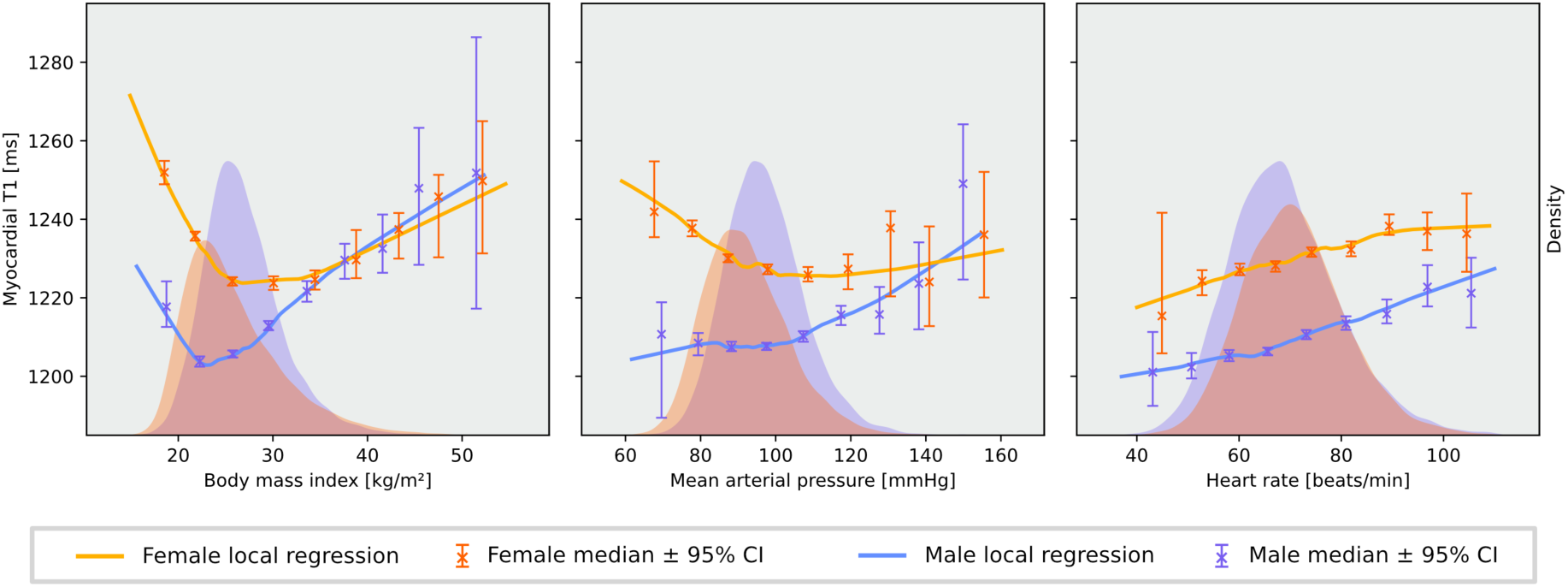
Local Regressions for Body Mass Index, Mean Arterial Pressure and Heart Rate. Locally estimated scatterplot smoothing, stratified by sex. Median and 95% confidence interval (bootstrapped with 1,000 iterations) are shown for nine equal-width bins. Values below the 0.1^st^ or above the 99.9^th^ percentile for heart rate were excluded. Two outliers were excluded for mean arterial pressure. Kernel density plots for men (blue) and women (orange) are displayed on a secondary y-axis along the same horizontal axis. CI: confidence interval

### Association of Hypertension Severity with Myocardial T1 by ESC Classification

In the sex-stratified, adjusted sub-analysis based on the ESC classification of measured blood pressure (Figure 6), a progressive association between hypertension severity and myocardial native T1 time was observed in men. Compared with normotensive individuals, T1 gradually increased in men from grade I (β = 3.05 [0.93–5.17] ms; *p* = 0.005) to grade III hypertension (β = 8.21 [2.40–14.02] ms; *p* = 0.006), while isolated diastolic or systolic hypertension showed no significant trends (*p* = 0.551 and *p* = 0.300, respectively). Among women, isolated diastolic hypertension (β = −3.89 [−7.27 to −0.52] ms; *p* = 0.024) and isolated systolic hypertension (β = −3.10 [−5.36 to −0.83] ms; *p* = 0.007) were significantly associated with lower myocardial T1 time, while no significant associations were observed for grade I (*p* = 0.236), grade II (*p* = 0.418), or grade III hypertension (*p* = 0.715).

**Figure 6:**
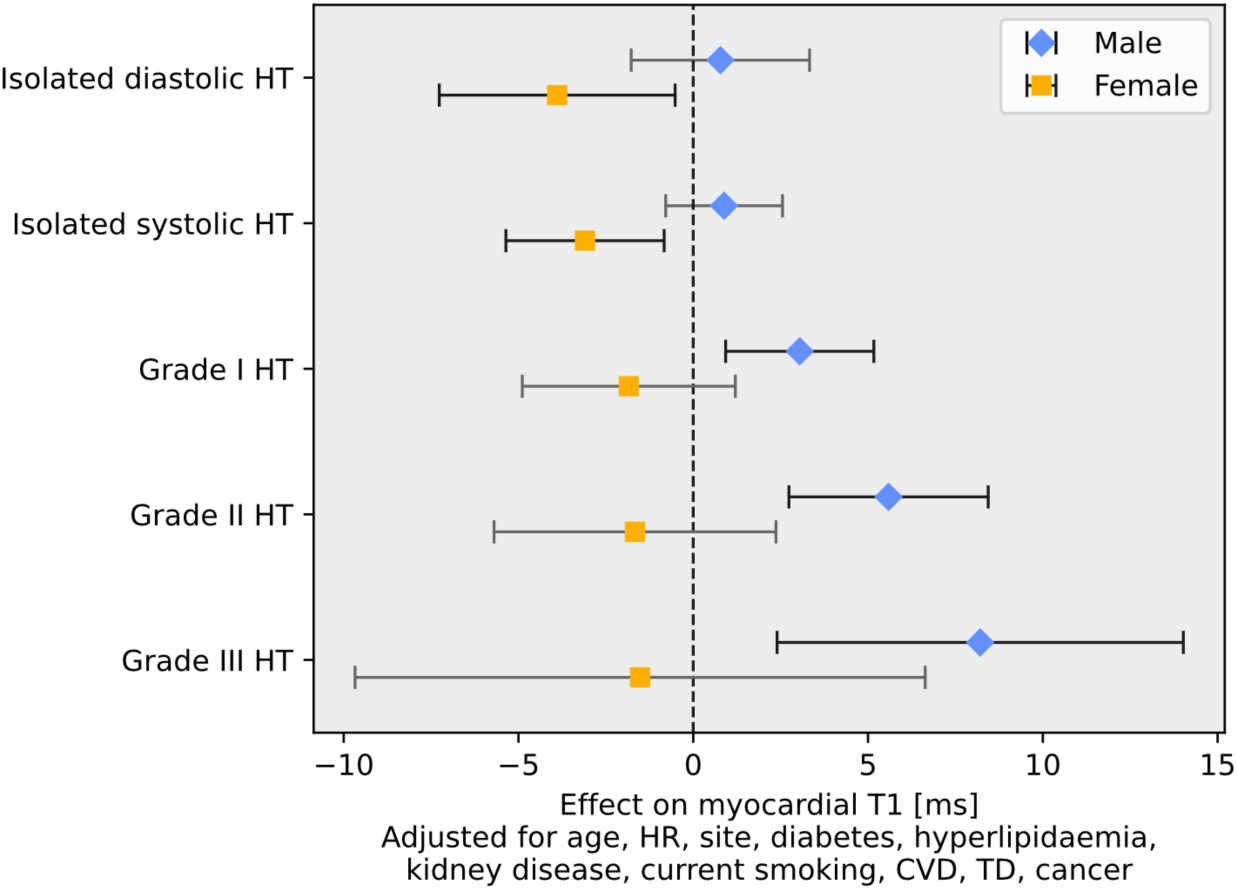
Sex-Stratified Associations of ESC Hypertension Categories with Myocardial T1. Unstandardized β coefficients and 95% confidence intervals from adjusted multiple linear regression, stratified by sex. Covariates included age, heart rate (HR), imaging site, diabetes, hyperlipidaemia, kidney disease, current smoking, cardiovascular disease (CVD), thyroid disease (TD), and cancer. Definitions for diabetes, hyperlipidaemia and kidney disease were based on medical history and clinical chemistry. HT: hypertension

## Discussion

### Summary of Key Findings

In this large-scale analysis of 3.0 T CMR-based T1 mapping in the NAKO study, we report associations between age, sex, and cardiometabolic risk factors with myocardial native T1 in 27,794 participants. Women exhibited overall significantly higher T1 values, with the sex difference progressively decreasing with age. Notably, we discovered an inverse relationship between hyperlipidaemia and myocardial T1 and found sex-specific associations for cardiometabolic risk factors, especially hypertension.

### Sex and Age Dependence of Myocardial T1

The observed sex differences in T1 are consistent with previous findings [13–15,25,26]. While the influence of age remains controversial in the literature [14,25], our analysis aligns with results from healthy participants in the UK Biobank [13] and results for men in the MESA study [26]. The broader age distribution in the NAKO cohort allowed inclusion of younger adults and the capture of the menopausal transition, which may contribute to the observed sex-age interactions. Although thinner myocardial walls in women [27] might increase sensitivity to partial volume effects, this alone is unlikely to account for the marked difference. The confirmatory analysis for septal segments further supports the internal validity of our findings. In the healthy subcohort, the age dependency was less pronounced and may therefore encompass differences in age-specific prevalence of diseases between sexes.

### Hyperlipidaemia and Myocardial T1

The significant association of elevated blood lipids with lower myocardial T1 is a novel and unexpected finding. Hyperlipidaemia typically promotes fibrosis via atherosclerotic pathways [28] and would therefore be more consistent with prolonged T1 times [1]. Our results suggest additional mechanisms, with possible explanations including altered myocardial lipid metabolism or early lipid infiltration. There is growing evidence that serum lipids have direct effects on the heart independent of atherosclerosis [12]. Obesity is associated with adipocyte infiltration of the myocardium [29] and although the underlying mechanisms may differ, lipid overload as in Fabry disease presents with shortened T1 times [30]. Another possible explanation would be a protective effect of statin therapy, which has been reported to attenuate extracellular volume expansion [31]. Lipid-lowering therapy showed a modest negative association with myocardial T1, suggesting a potential moderating role. However, this association did not reach statistical significance.

### Sex-Specific Association of Hypertension with Myocardial T1

The observation that hypertension is associated with elevated myocardial T1 in men and lower values in women, indicates a sex-specific cardiac response to pressure overload. One possible explanation lies in sex-specific hormonal and genetic regulation of blood pressure and target-organ damage [32]. The observed difference aligns with one study reporting greater strain impairment in hypertensive men [33], but contrasts with another [34], where female sex was independently associated with left ventricular remodelling. Differing results may be related to women losing the cardioprotective effects of oestrogen in the context of obesity and diabetes, potentially influencing the myocardial response to hypertension [32,33]. In men, the observed progressive increase in native T1 values across hypertension grades I to III suggests a dose-response relationship, where higher blood pressure levels are associated with greater myocardial remodelling.

### Diabetes, Kidney Disease, and Smoking

Our findings of elevated myocardial T1 in individuals with diabetes, kidney disease or current smoking likely reflect known pathophysiological mechanisms: In recent studies, diabetes was independently associated with higher native T1 values, indicating potential subclinical myocardial dysfunction before structural remodelling occurs [13,35]. Higher T1 values in individuals with diabetes may reflect expanded intracellular water volume in hypertrophied cardiomyocytes [35]. Additionally, advanced glycation end-products increasingly form in the presence of chronic hyperglycaemia and are linked to oxidative stress and coronary artery disease [36]. Diffuse myocardial fibrosis, as measured by myocardial native T1 and extracellular volume, has also been reported in early-stage chronic kidney disease [9] and is a recognized pathophysiological factor in haemodialysis patients [37,38]. Smoking is a well-established driver of systemic inflammation, oxidative stress, and atherosclerosis [11].

### Clinical Implications

Our findings demonstrate measurable myocardial changes associated with key components of the metabolic syndrome, suggesting that cardiometabolic comorbidities have subtle but detectable effects on the composition of myocardial tissue. Unlike previous studies that modelled T1 as a predictor of disease risk [13], our analysis considers T1 as the response variable to reflect structural sex-specific effects associated with cardiometabolic risk. The reported β coefficients thus represent average group-level shifts in tissue composition. Since not all individuals with a given risk factor show myocardial involvement, population heterogeneity dilutes mean group differences. However, even moderate effect sizes may reflect an increased prevalence of individuals with pathologic myocardial changes within exposed subgroups. To illustrate this, we performed a distributional analysis (Supplementary Figure 4) showing the proportion of participants with T1 values above or below one standard deviation from age- and sex-matched reference distributions. Even small mean differences correspond to a measurable enrichment in the distribution tails: for instance, 20.8% of participants with diabetes had T1 values above one standard deviation compared with 15.1% in healthy individuals. Recognizing myocardial tissue alterations in individuals with reversible cardiometabolic risk factor burden may facilitate the early identification of cardiac involvement and inform preventive strategies, even before the onset of symptomatic cardiovascular disease. At the same time, our analysis indicates that comorbidities such as diabetes or smoking do not limit the diagnostic capability of T1 mapping, as the magnitude of differences overall remains far below those seen in overt diseases such as amyloidosis or Fabry disease [30,39]. Finally, clinical interpretation of myocardial native T1 should consider sex-specific differences, as women show significantly higher baseline values.

### Limitations

Medical history in the NAKO cohort was collected via self-reported questionnaires, which may introduce recall or reporting error. To mitigate these limitations, self-reports were complemented with measured elevations in relevant blood biomarkers to reduce potential biases in the primary variables. While the population-based study design supports generalisability of the observed association patterns, absolute T1 values may not be directly transferable to different imaging settings or ethnically diverse cohorts.

## Conclusions

Our analysis provides large-scale population-based evidence linking cardiometabolic risk factors to myocardial native T1 time. Diabetes, kidney disease and smoking were associated with prolonged T1 times, indicating structural changes in myocardial tissue composition. Conversely, hyperlipidaemia was negatively associated with T1, requiring further investigation. We observed dynamic, age-dependent differences in T1 time between women and men, suggesting a strong influence of sex on myocardial composition, especially before the age of 50. While our findings for cardiometabolic risk factors underscore the complex interplay between metabolic and myocardial health, the overall moderate effect sizes and the inherent stability of native T1 mapping support its reliability as a diagnostic marker.

## Acknowledgements

This project was conducted with data (application numbers NAKO-744, NAKO-745) from the German National Cohort (NAKO). We thank all participants who took part in the NAKO study and the staff of this research initiative. We thank Siemens Healthineers, and especially Teodora Chitiboi, Jens Wetzl, and Christian Geppert, for providing the research segmentation algorithm.

## Data Availability

Data is available by application to the NAKO transfer office (https://transfer.nako.de).

## References

1. Messroghli DR, Moon JC, Ferreira VM, Grosse-Wortmann L, He T, Kellman P, et al. Clinical recommendations for cardiovascular magnetic resonance mapping of T1, T2, T2* and extracellular volume: A consensus statement by the Society for Cardiovascular Magnetic Resonance (SCMR) endorsed by the European Association for Cardiovascular Imaging (EACVI). J Cardiovasc Magn Reson. 2016;19:75. 10.1186/s12968-017-0389-8

2. Taylor AJ, Salerno M, Dharmakumar R, Jerosch-Herold M. T1 Mapping. JACC Cardiovasc Imaging. 2016;9:67–81. 10.1016/j.jcmg.2015.11.005

3. Schelbert EB, Messroghli DR. State of the Art: Clinical Applications of Cardiac T1 Mapping. Radiology. 2016;278:658–76. 10.1148/radiol.2016141802

4. Puntmann VO, D’Cruz D, Smith Z, Pastor A, Choong P, Voigt T, et al. Native Myocardial T1 Mapping by Cardiovascular Magnetic Resonance Imaging in Subclinical Cardiomyopathy in Patients With Systemic Lupus Erythematosus. Circ Cardiovasc Imaging. 2013;6:295–301. 10.1161/CIRCIMAGING.112.000151

5. Peters A, German National Cohort (NAKO) Consortium, Peters A, Greiser KH, Göttlicher S, Ahrens W, et al. Framework and baseline examination of the German National Cohort (NAKO). Eur J Epidemiol. 2022;37:1107–24. 10.1007/s10654-022-00890-5

6. Bamberg F, Schlett CL, Caspers S, Ringhof S, Günther M, Hirsch JG, et al. Baseline MRI Examination in the NAKO Health Study. Dtsch Arztebl Int. 2024;121:587–93. 10.3238/arztebl.m2024.0151

7. Pigeot I, Ahrens W. Epidemiology of metabolic syndrome. Pflüg Arch - Eur J Physiol. 2025;477:669–80. 10.1007/s00424-024-03051-7

8. Salvador DB, Gamba MR, Gonzalez-Jaramillo N, Gonzalez-Jaramillo V, Raguindin PFN, Minder B, et al. Diabetes and Myocardial Fibrosis. JACC Cardiovasc Imaging. 2022;15:796–808. 10.1016/j.jcmg.2021.12.008

9. Edwards NC, Moody WE, Yuan M, Hayer MK, Ferro CJ, Townend JN, et al. Diffuse Interstitial Fibrosis and Myocardial Dysfunction in Early Chronic Kidney Disease. Am J Cardiol. 2015;115:1311–7. 10.1016/j.amjcard.2015.02.015

10. González A, Ravassa S, López B, Moreno MU, Beaumont J, San José G, et al. Myocardial Remodeling in Hypertension: Toward a New View of Hypertensive Heart Disease. Hypertension. 2018;72:549–58. 10.1161/HYPERTENSIONAHA.118.11125

11. McEvoy JW, Blaha MJ, DeFilippis AP, Lima JAC, Bluemke DA, Hundley WG, et al. Cigarette Smoking and Cardiovascular Events: Role of Inflammation and Subclinical Atherosclerosis From the Multiethnic Study of Atherosclerosis. Arterioscler Thromb Vasc Biol. 2015;35:700–9. 10.1161/ATVBAHA.114.304562

12. Yao YS, Li TD, Zeng ZH. Mechanisms underlying direct actions of hyperlipidemia on myocardium: an updated review. Lipids Health Dis. 2020;19:23. 10.1186/s12944-019-1171-8

13. Raisi-Estabragh Z, McCracken C, Hann E, Condurache D-G, Harvey NC, Munroe PB, et al. Incident Clinical and Mortality Associations of Myocardial Native T1 in the UK Biobank. JACC Cardiovasc Imaging. 2023;16:450–60. 10.1016/j.jcmg.2022.06.011

14. Myhr KA, Andrés-Jensen L, Larsen BS, Kunkel JB, Kristensen CB, Vejlstrup N, et al. Sex- and age-related variations in myocardial tissue composition of the healthy heart: a native T1 mapping cohort study. Eur Heart J - Cardiovasc Imaging. 2024;25:1109–17. 10.1093/ehjci/jeae070

15. Cavus E, Schneider JN, Bei der Kellen R, di Carluccio E, Ziegler A, Tahir E, et al. Impact of Sex and Cardiovascular Risk Factors on Myocardial T1, Extracellular Volume Fraction, and T2 at 3 Tesla: Results From the Population-Based, Hamburg City Health Study. Circ Cardiovasc Imaging. American Heart Association; 2022;15:e014158. 10.1161/CIRCIMAGING.122.014158

16. Bamberg F, Kauczor H-U, Weckbach S, Schlett CL, Forsting M, Ladd SC, et al. Whole-Body MR Imaging in the German National Cohort: Rationale, Design, and Technical Background. Radiology. 2015;277:206–20. 10.1148/radiol.2015142272

17. Saad H, Ammann C, Hadler T, Bhoyroo Y, Reisdorf P, Veit J, et al. Assessing the robustness of an artificial intelligence segmentation model for quantitative cardiovascular magnetic resonance imaging across cardiac phenotypes. Int J Cardiovasc Imaging. 2025. 10.1007/s10554-025-03596-3

18. Popescu AB, Seitz A, Mahrholdt H, Wetzl J, Jacob A, Itu LM, et al. Deep learning-based segmentation of T1 and T2 cardiac MRI maps for automated disease detection [Preprint]. arXiv. 2025. 10.48550/ARXIV.2507.00903

19. American Heart Association Writing Group on Myocardial Segmentation and Registration for Cardiac Imaging: Cerqueira MD, Weissman NJ, Dilsizian V, Jacobs AK, Kaul S, et al. Standardized Myocardial Segmentation and Nomenclature for Tomographic Imaging of the Heart: A Statement for Healthcare Professionals From the Cardiac Imaging Committee of the Council on Clinical Cardiology of the American Heart Association. Circulation. 2002;105:539–42. 10.1161/hc0402.102975

20. Ammann C, Hadler T, Reisdorf P, Hickstein R, Noyan H, Gröschel J, et al. Cardiometry: Open-Source, Precise and Efficient Computation of Quantitative Parameters in Cardiovascular Magnetic Resonance [Abstract]. Proceedings of the ISMRM 33rd Annual Meeting and Exhibition. Honolulu, HI, USA: International Society for Magnetic Resonance in Medicine, Berkley, CA, USA; 2025. https://archive.ismrm.org/2025/0945.html

21. Kuss O, Becher H, Wienke A, Ittermann T, Ostrzinski S, Schipf S, et al. Statistical Analysis in the German National Cohort (NAKO) – Specific Aspects and General Recommendations. Eur J Epidemiol. 2022;37:429–36. 10.1007/s10654-022-00880-7

22. Kellman P, Hansen MS. T1-mapping in the heart: accuracy and precision. J Cardiovasc Magn Reson. 2014;16:2. 10.1186/1532-429X-16-2

23. Mach F, Baigent C, Catapano AL, Koskinas KC, Casula M, Badimon L, et al. 2019 ESC/EAS Guidelines for the management of dyslipidaemias: lipid modification to reduce cardiovascular risk. Eur Heart J. 2020;41:111–88. 10.1093/eurheartj/ehz455

24. McEvoy JW, McCarthy CP, Bruno RM, Brouwers S, Canavan MD, Ceconi C, et al. 2024 ESC Guidelines for the management of elevated blood pressure and hypertension. Eur Heart J. 2024;45:3912–4018. 10.1093/eurheartj/ehae178

25. Rosmini S, Bulluck H, Captur G, Treibel TA, Abdel-Gadir A, Bhuva AN, et al. Myocardial native T1 and extracellular volume with healthy ageing and gender. Eur Heart J - Cardiovasc Imaging. 2018;19:615–21. 10.1093/ehjci/jey034

26. Liu C-Y, Liu Y-C, Wu C, Armstrong A, Volpe GJ, Van Der Geest RJ, et al. Evaluation of Age-Related Interstitial Myocardial Fibrosis With Cardiac Magnetic Resonance Contrast-Enhanced T1 Mapping. J Am Coll Cardiol. 2013;62:1280–7. 10.1016/j.jacc.2013.05.078

27. Kawel-Boehm N, Hetzel SJ, Ambale-Venkatesh B, Captur G, Chin CWL, François CJ, et al. Society for Cardiovascular Magnetic Resonance reference values (“normal values”) in cardiovascular magnetic resonance: 2025 update. J Cardiovasc Magn Reson. 2025;27:101853. 10.1016/j.jocmr.2025.101853

28. Ross R, Harker L. Hyperlipidemia and Atherosclerosis: Chronic hyperlipidemia initiates and maintains lesions by endothelial cell desquamation and lipid accumulation. Science. 1976;193:1094–100. 10.1126/science.822515

29. Pathak RK, Mahajan R, Lau DH, Sanders P. The Implications of Obesity for Cardiac Arrhythmia Mechanisms and Management. Can J Cardiol. 2015;31:203–10. 10.1016/j.cjca.2014.10.027

30. Sado DM, White SK, Piechnik SK, Banypersad SM, Treibel T, Captur G, et al. Identification and Assessment of Anderson-Fabry Disease by Cardiovascular Magnetic Resonance Noncontrast Myocardial T1 Mapping. Circ Cardiovasc Imaging. 2013;6:392–8. 10.1161/CIRCIMAGING.112.000070

31. Juhasz V, Quinaglia T, Drobni ZD, Heemelaar JC, Neuberg DS, Han Y, et al. Atorvastatin and Myocardial Extracellular Volume Expansion During Anthracycline-Based Chemotherapy. JACC CardioOncology. 2025;7:125–37. 10.1016/j.jaccao.2024.11.008

32. Colafella KMM, Denton KM. Sex-specific differences in hypertension and associated cardiovascular disease. Nat Rev Nephrol. 2018;14:185–201. 10.1038/nrneph.2017.189

33. Tadic M, Cuspidi C, Celic V, Ivanovic B, Pencic B, Grassi G. The influence of sex on left ventricular strain in hypertensive population. J Hypertens. 2019;37:50–6. 10.1097/HJH.0000000000001838

34. Canciello G, Piccolo R, Izzo R, Bossone E, Pacella D, Lembo M, et al. Sex-Related Differences in Left Ventricular Geometry Patterns in Patients With Arterial Hypertension. JACC Adv. 2024;3:101256. 10.1016/j.jacadv.2024.101256

35. Lam B, Stromp TA, Hui Z, Vandsburger M. Myocardial native-T1 times are elevated as a function of hypertrophy, HbA1c, and heart rate in diabetic adults without diffuse fibrosis. Magn Reson Imaging. 2019;61:83–9. 10.1016/j.mri.2019.05.029

36. Fishman SL, Sonmez H, Basman C, Singh V, Poretsky L. The role of advanced glycation end-products in the development of coronary artery disease in patients with and without diabetes mellitus: a review. Mol Med. 2018;24:59. 10.1186/s10020-018-0060-3

37. Rutherford E, Talle MA, Mangion K, Bell E, Rauhalammi SM, Roditi G, et al. Defining myocardial tissue abnormalities in end-stage renal failure with cardiac magnetic resonance imaging using native T1 mapping. Kidney Int. 2016;90:845–52. 10.1016/j.kint.2016.06.014

38. Aoki J, Ikari Y, Nakajima H, Mori M, Sugimoto T, Hatori M, et al. Clinical and pathologic characteristics of dilated cardiomyopathy in hemodialysis patients. Kidney Int. 2005;67:333–40. 10.1111/j.1523-1755.2005.00086.x

39. Lavall D, Vosshage NH, Geßner R, Stöbe S, Ebel S, Denecke T, et al. Native T1 mapping for the diagnosis of cardiac amyloidosis in patients with left ventricular hypertrophy. Clin Res Cardiol. 2023;112:334–42. 10.1007/s00392-022-02005-2

