## Supplementary Material for "Myocardial Native T1 Mapping in the German National Cohort (NAKO): Associations with Age, Sex, and Cardiometabolic Risk Factors"

*Supplementary Table 1: Septal Segments – Mean Myocardial T1 for Age Groups by Sex*

| Age | Male | | Female | |
|---|---|---|---|---|
| Group | mean ± SD | n | mean ± SD | n |
| 20-29 | 1,223 ± 30 | 1,532 | 1,249 ± 35 | 1,166 |
| 30-39 | 1,225 ± 33 | 2,082 | 1,253 ± 36 | 1,426 |
| 40-49 | 1,227 ± 35 | 4,237 | 1,252 ± 36 | 3,443 |
| 50-59 | 1,229 ± 39 | 4,341 | 1,245 ± 35 | 3,636 |
| 60-75 | 1,232 ± 42 | 3,181 | 1,241 ± 35 | 2,714 |
| Total | 1,228 ± 37 | 15,372 | 1,247 ± 36 | 12,385 |

SD: standard deviation; n: number of subjects

*Supplementary Table 2: Beta Coefficients and Confidence Intervals from Linear Regressions*

| | Cohort | | Male | | Female | |
|---|---|---|---|---|---|---|
| | β [95% CI] | *p* | β [95% CI] | *p* | β [95% CI] | *p* |
| Diabetes or HbA1c ≥ 48 mmol/mol | 4.14 [2.23–6.06] | < 0.001 | 6.48 [3.92–9.05] | < 0.001 | 1.64 [-1.23 to 4.52] | 0.263 |
| Hyperlipidaemia or LDL-c. ≥ 3.0 mmol/l | -4.69 [-5.59 to -3.79] | < 0.001 | -3.67 [-4.92 to -2.43] | < 0.001 | -5.59 [-6.90 to -4.29] | < 0.001 |
| Hypertension or BP ≥ 140/90 mmHg | -0.04 [-0.94 to 0.86] | 0.931 | 2.50 [1.31–3.68] | < 0.001 | -3.83 [-5.22 to -2.44] | < 0.001 |
| Kidney disease or crea. ≥ 106/88 µmol/l | 3.64 [1.63–5.64] | < 0.001 | 5.06 [2.42–7.70] | < 0.001 | 1.94 [-1.13 to 5.01] | 0.215 |
| Current smoker | 6.60 [5.59–7.62] | < 0.001 | 7.97 [6.62–9.33] | < 0.001 | 4.63 [3.11–6.15] | < 0.001 |
| Myocardial infarction | 11.40 [3.37–19.43] | 0.005 | 7.43 [-1.84 to 16.70] | 0.116 | 19.51 [2.66–36.37] | 0.023 |
| CAD or angina pectoris | 1.82 [-3.01 to 6.66] | 0.460 | 1.86 [-4.00 to 7.72] | 0.535 | 1.06 [-7.61 to 9.74] | 0.810 |
| Heart failure | 5.85 [2.25–9.46] | 0.001 | 10.81 [5.92–15.69] | < 0.001 | -0.21 [-5.56 to 5.13] | 0.937 |
| Arrhythmia | 3.16 [1.56–4.76] | < 0.001 | 4.24 [1.96–6.52] | < 0.001 | 1.95 [-0.30 to 4.19] | 0.089 |
| Peripheral artery disease | 2.87 [-1.47 to 7.20] | 0.195 | 5.24 [-0.47 to 10.94] | 0.072 | -0.62 [-7.27 to 6.03] | 0.855 |
| Thyroid disease | -0.19 [-1.26 to 0.87] | 0.720 | 1.27 [-0.65 to 3.19] | 0.194 | -0.62 [-1.88 to 0.64] | 0.334 |
| Cancer | -0.39 [-2.20 to 1.42] | 0.672 | -0.13 [-2.85 to 2.58] | 0.923 | -0.50 [-2.90 to 1.90] | 0.681 |

Unstandardized β coefficients from multiple linear regression models, adjusted for age, heart rate, imaging site as well as sex and age×sex if applicable. Physician-diagnosed conditions were collected by a standardized personal interview. LDL-c: LDL-cholesterol; BP: blood pressure; crea: serum creatinine; CAD: coronary artery disease; CI: confidence interval

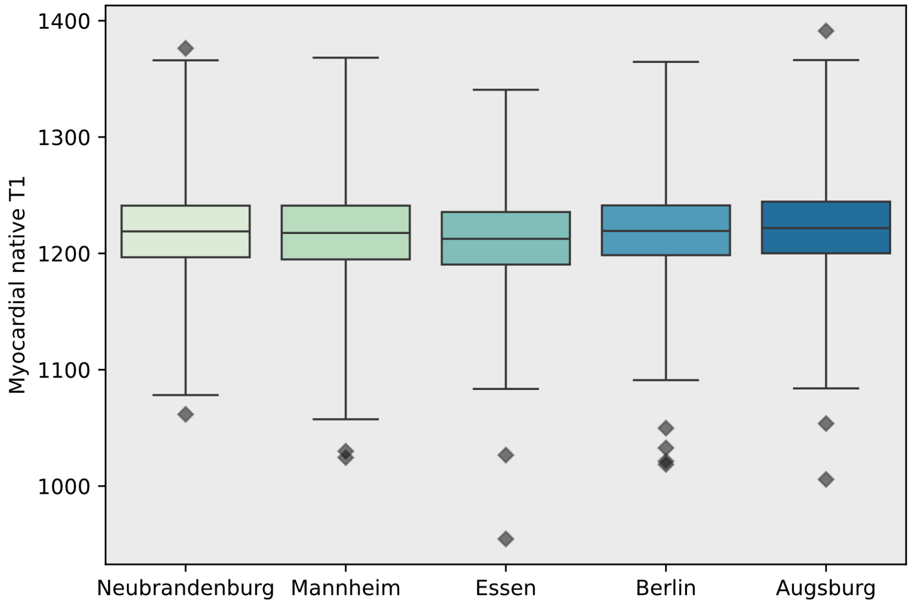


*Supplementary Figure 1: Boxplots for Inter-Site Differences in Myocardial Native T1*

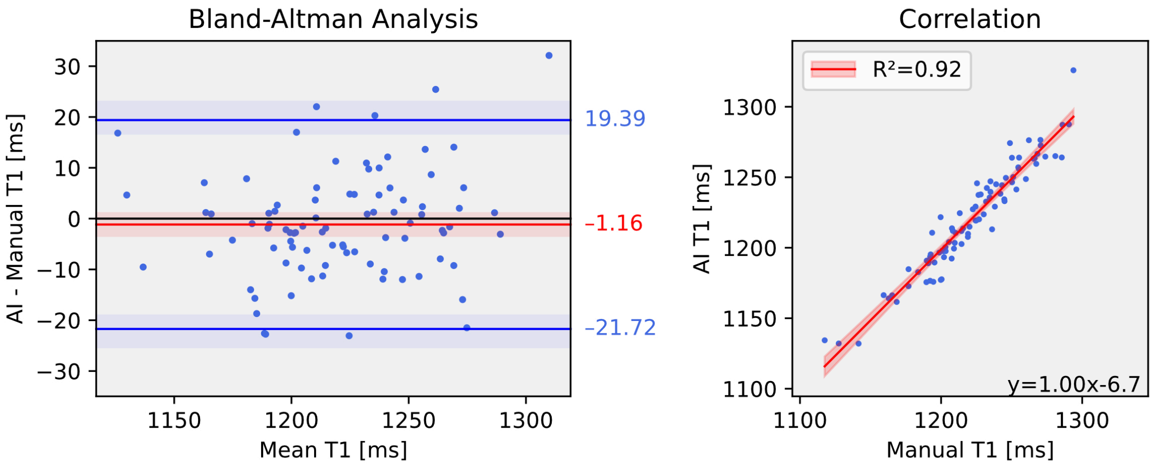


*Supplementary Figure 2: Bland-Altman and Correlation Plots for the Manual Validation Sample*

AI: artificial intelligence

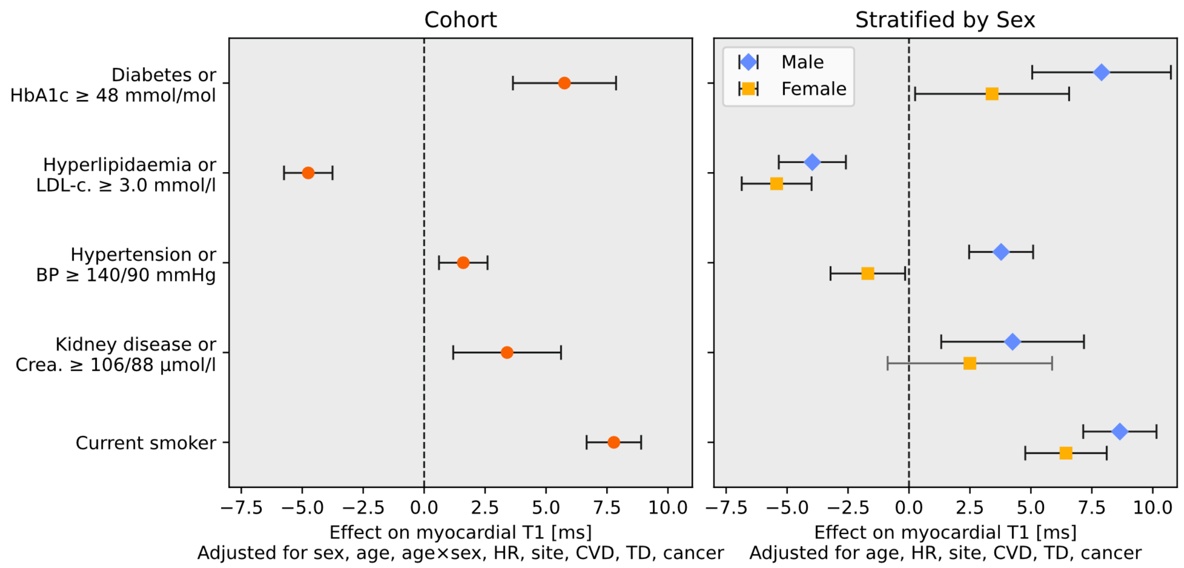


*Supplementary Figure 3: Septal Segments – Associations of Cardiometabolic Risk Factors with Myocardial T1*

Displayed are unstandardized β coefficients and 95% confidence intervals from linear regression models for the whole sample (left panel) and stratified by sex (right panel). Covariates included sex, age, age×sex, heart rate (HR), imaging site, cardiovascular disease (CVD), thyroid disease (TD), and cancer. LDL-c.: LDL-cholesterol; BP: blood pressure; Crea.: serum creatinine

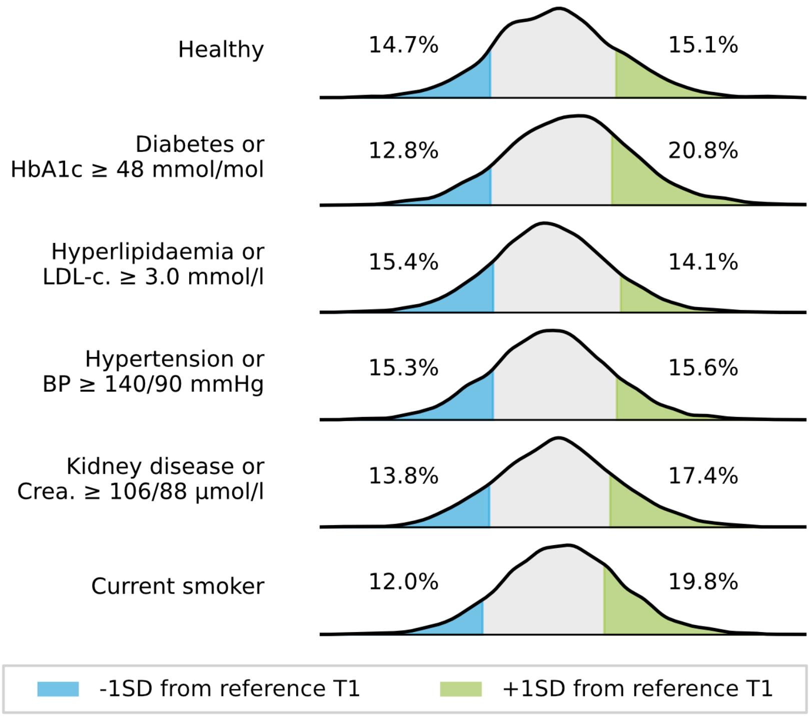


*Supplementary Figure 4: Analysis of Myocardial T1 Distribution Tails*

Density plots for myocardial native T1 in healthy and cardiometabolic risk groups. Shaded areas indicate the proportion of individuals with myocardial T1 values above or below one standard deviation (SD) from age- (10-year groups) and sex-matched reference distributions in the overall cohort. Calculated cutoff values (mean ± 1SD) vary depending on age and sex but are shown in simplified form. LDL-c: LDL-cholesterol; BP: blood pressure; Crea.: serum creatinine
